# Shortened SARS-CoV-2 shedding in saliva during early Omicron compared to wild-type pandemic phase

**DOI:** 10.1101/2024.09.11.24313457

**Authors:** Eva Kozanli, Angelique M.A.M. Winkel, Alvin X. Han, Sharon van den Brink, Annemarie van den Brandt, Milly E. Haverkort, Sjoerd Euser, Colin A. Russell, Menno D. de Jong, Marlies A. van Houten, Steven F.L. van Lelyveld, Dirk Eggink

## Abstract

This study compared the dynamics of SARS-CoV-2 viral shedding in saliva between wild-type virus-infected and Omicron-infected household cohorts. Pre-existing immunity in participants likely shortens duration of viral shedding and lowers viral load peaks. Dense saliva sampling can be a convenient tool to study viral load dynamics.

## Introduction

SARS-CoV-2 has caused a pandemic with a large number of respiratory infections worldwide. Studying the viral load dynamics of SARS-CoV-2 infections is helpful for understanding pathogenesis and transmission of the virus. Viral load dynamics can be influenced by various clinical host-factors, the emergence of novel variants and immunization (1).

SARS-CoV-2 infections are conventionally detected performing reverse transcriptase polymerase chain reaction (RT-PCR) on nasopharyngeal/oropharyngeal swabs (NP/OP). The cycle threshold (Ct) value of the PCR reaction cannot be directly translated to infectious virus particles (1), but nevertheless provides a valuable marker for viral load and thereby potentially for severity of infection and transmissibility. To date, NP/OP swabs is the gold standard for respiratory diagnostics (2), but since saliva can be self-sampled, it offers a convenient diagnostic method (3). Additionally, it is easier to facilitate longitudinal sampling of infected individuals and is especially beneficial for studying shedding in children, as it is non-invasive. Previous studies have found high sensitivity for SARS-CoV-2 detection in saliva compared to NP/OP swabs (4). Therefore, we hypothesize that shedding dynamics and underlying covariates could be effectively studied in saliva.

To our best knowledge, data from longitudinal household studies using saliva to characterize viral shedding dynamics is limited. We conducted two identical household studies during two different pandemic phases: SARSLIVA1 (SL1) during the circulation of wild-type (ancestral) SARS-CoV-2 in 2020 (3); and SARSLIVA2 (SL2) during circulation of Omicron variants in 2022 in a population with pre-existing immunity(5). Thus, we aim to investigate shedding dynamics and impact of SARS-CoV-2 variants in saliva, and the role of host-immune status.

## Methods

### Participants and sampling

Two identical prospective household cohort studies were performed, named SARSLIVA 1 (SL1) and SARSLIVA 2 (SL2). SL1 took place during the circulation of wild-type SARS-CoV-2 from October to December 2020 (3); and SL2 during circulation of Omicron variant, mainly BA.2, in March and April 2022 in a population with pre-existing immunity (5). Participants were recruited through community testing in the Netherlands as described (3). If these individuals (index cases) tested positive, were younger than 65 years old and had at least two other household members were willing to participate, their households were included. All participants completed questionnaires regarding medical history and updated their health status throughout the study. Dense interval self-sampling of saliva led to 10 samples in 42 days. Capillary serum samples were collected at day 1 and day 42 for SL2 and at day 42 for SL1. Index cases also indicated their date of symptom onset.

Written informed consent was obtained from all participants. This study was reviewed and approved by the Medical Ethical Committee of the Amsterdam University Medical Centre, The Netherlands (reference number 2020.436 (SARSLIVA 1) and 2022.0073 (SARSLIVA 2.0)).

### Outcomes and definitions

Primary outcomes are the viral load peak and viral shedding duration. The peak is defined as the minimum Ct-values observed during the testing period. Duration is defined as the number of days between the first positive and the midpoint between the last positive PCR result and first negative result. The last positive PCR result should be followed by at least two negative PCR results. Secondary outcomes are determinants that influence the viral load peak and shedding duration, which include sex, age, disease severity, weight class and pre- and post-infection antibody levels.

### Laboratory assays: RT-PCR and serology

RT-PCR for SARS-CoV-2 was performed on each self-sampled saliva specimen (6). Ct-values under 40 were considered SARS-CoV-2 positive. Sera were analyzed for presence of SARS-COV-2 specific antibodies by protein microarray for the spike protein, using dose-response curves from serial dilutions to determine fluorescent signal (7).

### Cohort comparisons

Bayes t-tests were performed to compare the mean viral load peak and mean viral shedding duration between cohorts. We assumed that there was strong evidence to support the alternate hypothesis (i.e. differences between cohorts) if the Bayes Factor (BF) exceeded 10. Sensitivity analyses were performed for our index cases only to correct for enrollment time by including interval between symptom onset and inclusion.

### Bayesian linear regression for viral load decline

We used a Bayesian hierarchical linear regression model to estimate rates of viral load decline (8). We included all participants whose symptom onset dates were known. We estimated the response variable difference in Ct-value (Y) over time since symptom onset (t) for each sample (s):

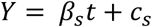

The full model details, including priors used, are described in Supplementary methods.

### Bayesian hierarchical ANOVA model for shedding duration and minimum Ct-value

An Bayesian hierarchical ANOVA model was used to estimate effects of covariates on the viral load peak and shedding duration, separately for SL1 and SL2. Full model and prior details are described in Supplementary methods.

## Results

### Participants and sampling

The SL1 and SL2 cohort yielded 213 and 130 SARS-CoV-2 positive participants, of which 80 and 65 index cases, from 85 and 69 households, respectively (Supplemental table 1). In total, 3273 saliva samples were obtained, with an average of 9.5 of the 10 samples per participant included in our analyses. Of SL2 participants, 73.2% were vaccinated (at least once) at inclusion. None of our participants was hospitalized.

### Viral load and shedding duration in different pandemic phases

We observed a difference in inclusion time (i.e. time from NP/OP swab to first day of saliva sampling) between SL1 (median: 5 days) and SL2 (median: 3 days, BF > 100), which was corrected for in our sensitivity analysis, reported as the corrected mean.

Shedding duration during SL1 (mean 25 days; corrected mean 29 days) was significantly longer as compared to SL2 (mean 15 days; corrected mean 17 days, initial and corrected BF>100). Minimum Ct-value (mean SL1: 26.3; SL2: 25.6; corrected mean: SL1: 26.2; SL2: 25.6), or viral load peak, were comparable between the two cohorts, and thus during these two stages of the pandemic. Moreover, modeling the decrease in viral load over time (Figure 1a/b), revealed a faster decline of viral load for SL2 than for SL1 (slope Ct-value per day SL1: 0.266; and SL2: 0.491).

**Figure 1.**
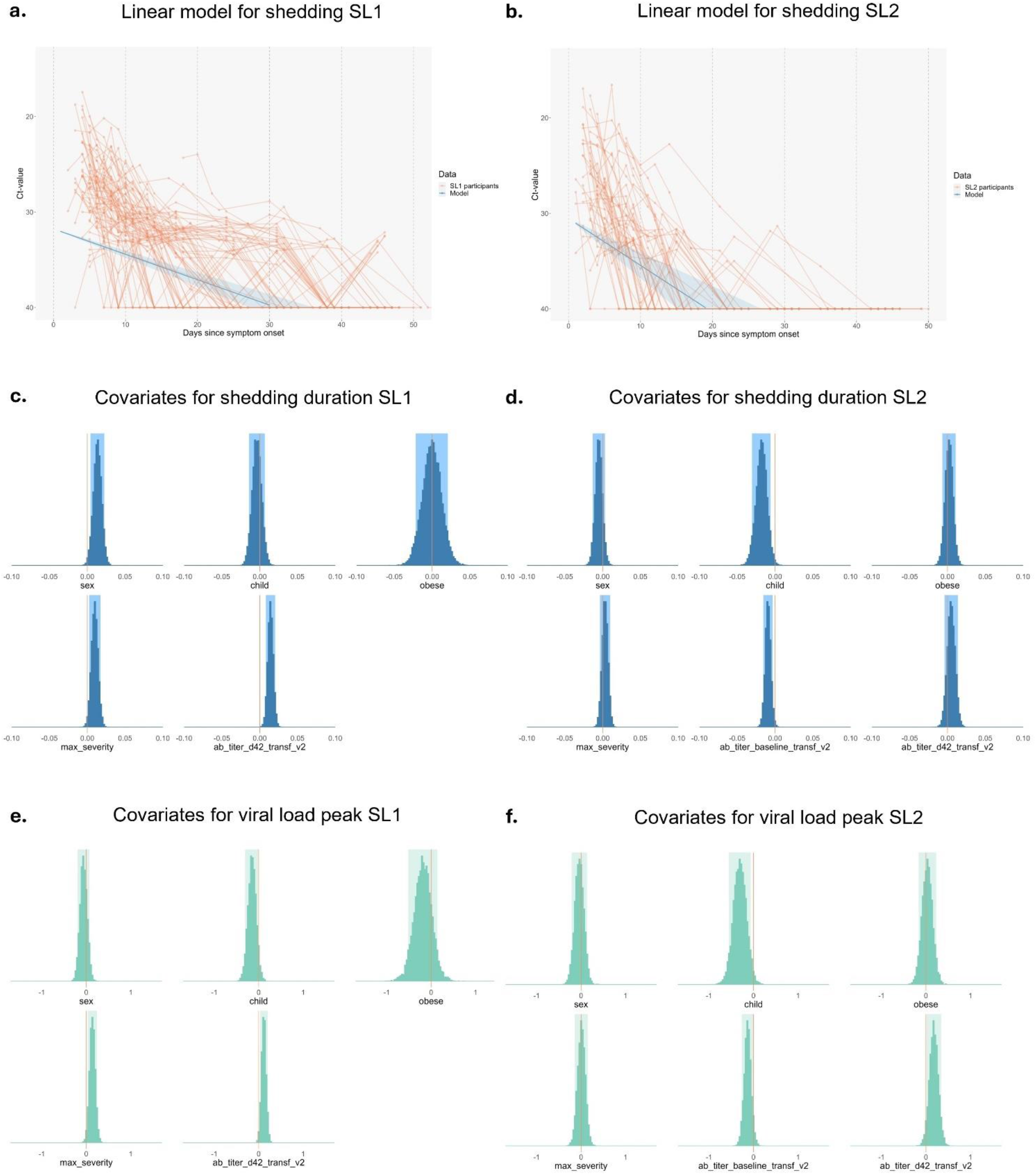
Viral shedding dynamics for two cohorts. **(a/b)** Bayesian model shows the linear decrease of viral load (increase of Ct-value) over time using inverted Ct-value on the y-axis. The two models for SL1 (a) and SL2 (b) show a steeper decline for SL2. **(c/d)** Visualization of covariates included that might influence duration of shedding for the two different cohorts, based on a Bayesian ANOVA model. SL1 (c) shows that men have prolonged duration of shedding and antibody titers at day 42 are higher when shedding is longer. In SL2 (d) high baseline titer affects shedding duration and children shed shorter. **(e/f)** Models show the effect size of covariates when comparing the minimum Ct-value (viral load peak) in the different cohorts. In both cohorts, children had lower viral load than adults. Also, antibodies were higher at day 42 in participants with high viral loads. We found that increase in disease severity correlated with higher peak viral load peak in SL1, but not in SL2 (f). In SL2 higher baseline antibody titers were associated with lower viral loads.

### Covariates for shedding duration and viral load peak

We analyzed the adjusted effects of sex, age, weight class, disease severity and pre- and post-infection immunity on shedding duration (Figure 1c/1d) and viral load peak (Figure 1e/1f). During SL1, males were more likely to shed virus longer than females (Figure 1c). Disease severity played a role in this early phase, as more severe disease showed a significant association with longer shedding duration and higher viral loads in SL1. In SL2, children shed shorter and had lower peak viral loads compared to adults. Participants in both SL1 as SL2 with high viral loads developed higher SARS-CoV-2 specific antibody titers 42 days post-infection. In SL1, longer duration of shedding was also associated with high antibody titers post-infection. Additionally, in SL2 we found that high baseline titers led to lower viral load and shorter shedding (Figure 1 c-f, Supplementary Table 2).

## Discussion

We used dense saliva sampling to compare SARS-CoV-2 shedding dynamics between mildly infected household cohorts from the 2020, wild-type SARS-CoV-2 phase of the pandemic (SL1) and the 2022 Omicron (post-immunization) phase (SL2). Duration of shedding of SARS-CoV-2 was shorter during SL2, but viral load peak was similar. Children in the SL2 cohort experienced lower viral loads and shorter shedding periods than adults, independent of covariates. Furthermore, we found that SARS-CoV-2 specific antibodies are protective against high viral loads and longer duration of shedding in saliva.

Viral shedding dynamics differed in pandemic phases. It is difficult to distinguish between the effect of immunity and the intrinsic characteristics of the circulating variant, since the two are strongly correlated throughout time and inevitably intertwined. However, the presence of higher antibody titers in participants with low viral loads suggests a protective contribution of immunity, which is consistent with findings from other studies involving vaccinated participants (9). In contrast, the viral load peak was similar in our two cohorts, despite pre-existing immunity by vaccination or earlier confirmed infection for most participants in SL2, and no pre-existing immunity in SL1. We hypothesize that the protective effect of pre-existing immunity against infection and severe disease is counteracted in the infected participants by properties of the Omicron variant, leading to a comparable shedding peak in both cohorts. It is often suggested that high viral loads are associated with disease severity. Our study confirms this increased severity correlated to viral load for SL1. However, for SL2 we do not see this same effect, so despite similar peak viral load, no correlation is seen with disease severity in SL2, likely attributable to both host immunity and characteristics of Omicron variant.

This study showed that shedding duration in both cohorts was much longer than isolation advices stated over the course of the pandemic. During the pandemic, there has been debate about the correlation between shedding and viral transmission, partly because the detection of RNA does not directly translate to the detection of infectious virus (1). Our study showed that children have lower viral loads and shed shorter than adults during the omicron phase. However, in the previous study that investigated transmission within the SL2 cohort (5), children contributed largely to transmission within households. This suggests that transmission is multifactorial, involving presence of immunity and behavioral factors. Consequently, we emphasize that transmission cannot be predicted solely by shedding dynamics, which should be considered in policy making based on these shedding dynamics.

A limitation of this study was posed by the fluctuating Ct-values in saliva samples which affected the resolution of viral load curves. Nonetheless, we show that saliva is a reliable measure for viral load, as has been suggested before (1, 10, 11). Moreover, we observed a longer inclusion time in SL1 than SL2, due to delayed reporting of results and limited accessibility of testing in the first pandemic phase. However, we corrected for this difference in our sensitivity analysis, which showed that inclusion time did not affect the outcome on shedding duration. Although this study is not the first to compare viral loads of different variants(12), this study is the first to use dense, saliva sampling over a period of six weeks to compare shedding.

To our best knowledge, we here describe a unique approach by analyzing saliva to compare viral shedding dynamics in different stages of the COVID-19 pandemic using two household cohorts separated in time but with identical study designs. Saliva specimens provide a convenient, non-invasive and adaptable method to study the effect of SARS-CoV-2 variant and immunity on shedding dynamics, which might contribute to future early surveillance for epidemic outbreaks.

## Data Availability

De-identified data collected for the study, including individual participant data collected during the study and a data dictionary, will be shared upon reasonable request. These requests will be discussed with all project partners (Spaarne Gasthuis, Public Health Service Kennemerland and RIVM). Requests should be directed to. These requests will be reviewed and approved by the investigator and project partners based on scientific merit. To gain access, data requesters will need to sign a data access agreement. Privacy-sensitive data, which is traceable to the participant, will not be shared.

## DECLARATIONS

### Declaration of interests

We declare no competing interests.

## Funding

The SARSLIVA 1 study was supported by internal funds from the National Institute for Public Health and the Environment (RIVM), and by The Netherlands Organization for Health Research and Development (ZonMw), grant number 10430012010017, financed in part by The Netherlands Ministry of Health, Welfare and Sport. E. A. M. S. reports employment with RIVM, who supported the research for this work. Moreover, both SARSLIVA studies were supported by internal funds from Spaarne Gasthuis.

## Authors’ contributions

DE, SvL, MAvH, MH, SE, EK, AW were responsible for the conceptualization and design of the study and participated in the acquisition of data; DE, AvdB, SvdB coordinated and performed laboratory analyses; EK and AXH designed the model and analyzed the data. EK, AW, AXH, DE, SvL, MAvH, were responsible for interpretation of the analyses and verified the underlying data; AW and EK wrote the manuscript with review and editing put from all authors. DE, SvL, MAvH, CAR, MDdJ were responsible for resources, supervision, project administration and funding acquisition. All authors had full access to all the data and reviewed and approved the final version of the manuscript.

## Acknowledgements

We thank all participants of SARSLIVA 1 and 2 without whom this study could not have been possible. We also thank the team of the Public Health Services (GGD) Kennemerland for providing information to the (possible) participants, the laboratory team of the National Institute for Public Health and the Environment (RIVM), including Gert-Jan Godeke, Bas van der Veer, Jordy de Bakker, Afke Vogelzang, Jeroen Cremer, Lisa Wijsman, Ryanne Jaarsma, Kim Freriks, Lynn Aarts, Sanne Bos, Mansoer Elahi, Jil Kocken for the laboratory analyses and the research team of the Spaarne Gasthuis Academy, particularly Lisa Kolodziej, Jordy Koole, Jacqueline Zonneveld, Sandra Kaamer van Hoegee, Mara van Roermund, Josseline Veldhuijzen and Yara Sijm.

## Notes

### Competing Interest Statement

The authors have declared no competing interest.

