## Supplementary Material for "Shortened SARS-CoV-2 shedding in saliva during early Omicron compared to wild-type pandemic phase"

### Supplementary text

#### Supplementary Methods

##### Type 1: Bayesian Hierarchical linear regression for shedding dynamics

In order to model the decline in viral load over time, we set up a Bayesian model for the differences in Ct value for the two different cohorts. We included all participants of which the symptom onset date is known. We used a single phase model using following formula to estimate the response variable difference in Ct value (Y) over time since symptom onset (t) for each sample (s), as has been described previously (1).

$$Y = \beta_s t + c_s$$

We assumed a prior distribution for each of our parameters. We placed a Gaussian prior on the intercept. The internal variation between samples are taken into account by assuming a normal distribution of the sample specific effect size.

$$c \sim normal(0,1)$$

$$\beta_s \sim normal(\beta, \sigma_\beta)$$

For the mean of the prior of the effect size, we placed a weakly informative Student T distribution as prior and assigned a half normal standard deviation.

$$\beta \sim T(3, 0, 2.5)$$

$$\sigma_\beta \sim half - normal(0, 1)$$

We assumed our response variable Y to be distributed according to a student-T distribution.

$$Y \sim T(\nu_y, Y, \sigma_y)$$

We assumed priors of the parameters of this distribution:

$$\nu \sim exponential\left(\frac{1}{30}\right)$$

$$\sigma_y \sim cauchy(0, 5)$$

We used a normal distribution (mean =0 and variance = 0.5) for offset values for each distribution to train our models.

#### Covariates and transformations

We included the covariates sex, age, disease severity, weight class, baseline and day 42 antibody titer. Sex was defined as male or female. Age is split up in the categories child (under 18 years) and adults (18 years and above) at inclusion. Disease severity for SARS-CoV-2 infection is classified as severity score one to four, as described earlier (2, 3): (1) no symptoms related to respiratory infection, (2) mild symptoms (pharyngitis, rhinitis, mild dyspnea, mild or moderate coughing, gustatory dysfunction or olfactory dysfunction), (3) moderate symptoms (moderate or severe dyspnea, severe coughing, temperature > 38°C or a pneumonia diagnosed by a physician), and (4) hospital admission due to SARS-CoV-2-related symptoms. Weight class was defined as obese or not obese, based on BMI score. Antibody titers refer to SARS-CoV-2 Spike specific antigens tested at inclusion of study (baseline) and approximately 42 days after inclusion, which are categorized into (1) . Other covariates like number/type of vaccines, comorbidities, smoking habits and education level have been considered, but groups were too small to further investigate these.

All variables are transformed around the mean or categorized in order to fit the model.

#### Type 2: Bayesian Hierarchical ANOVA model for minimum Ct value and for shedding duration

A Bayesian Hierarchical ANOVA model was used to estimate the effect of covariates on the minimum Ct value of each participant. Each participant had up to possibly 10 saliva samples in an interval of 42 days, of which Ct values where determined. The lowest value out of these 10 samples is indicated as the minimum Ct value which corresponds with the peak of viral load.

A similar model was applied to the shedding duration, which is defined as the first day where a Ct value under 40 was measured, to the last day (i.e. the mean of two sequential sampling days) from symptom or shedding onset.

For both models we look at the effect size (β), which is estimated for various covariates (X) as mentioned above to infer the correlation between these and minimum Ct value. The formula for this model is:

$$Y = c_s + \sum_k \beta_{s,k} X_k$$

In this model, Y and its parameters follow the same distribution as explained before. In addition, we look at participant specific effect sizes of each variable in our model and assume a normal distribution of the sample specific effect size. Similar distributions apply to the priors in this model. Again, estimation of the effect size follows a normal distribution:

$$\beta_{s,k} \sim normal(\beta_k, \sigma_{\beta_k})$$

The prior for $\sigma_y$ has been narrowed for the duration model to the following:

$$\sigma_y \sim cauchy(0, 2)$$

And for the hierarchy in our model, we, again, assumed our response variable Y to be distributed according to a student-T distribution.

$$Y \sim T(\nu_y, Y, \sigma_y)$$

**Supplementary Table 1: Baseline tables.** Table shows baseline characteristics for both cohorts. Looking at all individuals (**a**) and index cases only (**b**).

| **Table 1a: Baseline characteristics all participants** | | | |
|---|---|---|---|
| **Characteristic** | **Overall**, N = 343[1] | **Study** | |
| | | **Sarsliva 1.0**, N = 213[1] | **Sarsliva 2.0**, N = 130[1] |
| **Sex** | | | |
| Male | 166 (48%) | 110 (52%) | 56 (43%) |
| Female | 177 (52%) | 103 (48%) | 74 (57%) |
| **Age, years** | 34 (13-46) | 30 (15-46) | 36 (10-47) |
| **Child/adult** | | | |
| Adult (18 years and older) | 217 (63%) | 136 (64%) | 81 (62%) |
| Child (under 18 years old) | 126 (37%) | 77 (36%) | 49 (38%) |
| **Age group** | | | |
| 0 to 11 years old | 69 (20%) | 33 (15%) | 36 (28%) |
| 12 to 17 years old | 57 (17%) | 44 (21%) | 13 (10%) |
| 18 to 39 years old | 79 (23%) | 55 (26%) | 24 (18%) |
| 40-49 years old | 84 (24%) | 47 (22%) | 37 (28%) |
| 50 years and older | 54 (16%) | 34 (16%) | 20 (15%) |
| **Disease severity** | | | |
| Asymptomatic | 62 (18%) | 50 (23%) | 12 (9.2%) |
| Mild symptoms | 206 (60%) | 122 (57%) | 84 (65%) |
| Moderately severe symptoms | 74 (22%) | 40 (19%) | 34 (26%) |
| Severe symptoms | 1 (0.3%) | 1 (0.5%) | 0 (0%) |
| **Weight** [2] | | | |
| Obesity | 34 (9.9%) | 17 (8.0%) | 17 (13%) |
| No obesity | 308 (90%) | 195 (91%) | 113 (86%) |
| Unknown | 1 | 1 | 0 |
| **Any comorbidities** | 117 (34%) | 61 (29%) | 56 (43%) |
| **Atopy** | 94 (28%) | 47 (23%) | 47 (36%) |
| Unknown | 6 | 6 | 0 |
| Cardiac comorbidities | 4 (1.2%) | 2 (0.9%) | 2 (1.5%) |
| Pulmonary comorbidities | 3 (0.9%) | 2 (0.9%) | 1 (0.8%) |
| Immunological comorbidities | 4 (1.2%) | 1 (0.5%) | 3 (2.3%) |
| Diabetes Mellitus | 4 (1.2%) | 2 (0.9%) | 2 (1.5%) |
| Rheumatologi**c** comorbidities | 2 (0.6%) | 1 (0.5%) | 1 (0.8%) |
| Other comorbidities | 25 (7.3%) | 14 (6.6%) | 11 (8.5%) |
| **Smoking** | 8 (2.3%) | 5 (2.3%) | 3 (2.3%) |
| **Immunity status** | | | |
| No immunity | 235 (69%) | 213 (100%) | 22 (17%) |
| Previous infection | 13 (3.8%) | 0 (0%) | 13 (10%) |
| Vaccination | 83 (24%) | 0 (0%) | 83 (64%) |
| Hybrid immunity [3] | 12 (3.5%) | 0 (0%) | 12 (9.2%) |
| **Antibody titer day 1** | | | |
| **<10** | 10 (8.6%) | - | 10 (8.6%) |
| **10-100** | 2 (1.7%) | - | 2 (1.7%) |
| **100-1000** | 12 (10%) | - | 12 (10%) |
| **1000-10.000** | 66 (57%) | - | 66 (57%) |
| **>10.000** | 26 (22%) | - | 26 (22%) |

| | | | |
|---|---|---|---|
| Unknown | 227 | 213 | 14 |
| **Antibody titer day 42** | | | |
| **<10** | 14 (4.6%) | 13 (6.8%) | 1 (0.9%) |
| **10-100** | 12 (3.9%) | 12 (6.3%) | 0 (0%) |
| **100-1000** | 106 (35%) | 96 (51%) | 10 (8.7%) |
| **1000-10.000** | 102 (33%) | 69 (36%) | 33 (29%) |
| **>10.000** | 71 (23%) | 0 (0%) | 71 (62%) |
| Unknown | 38 | 23 | 15 |
| **Time from inclusion to last vaccination (days)** | 88 (72-114) | - | 88 (72-114) |
| Unknown | 248 | 213 | 35 |
| **Time from inclusion to last infection (days)** | 115 (96-321) | - | 115 (96-321) |
| Unknown | 318 | 213 | 105 |
| **Time from inclusion to last vaccination or exposure (days)** | 89 (73-113) | - | 89 (73-113) |
| Unknown | 235 | 213 | 22 |

[1] n (%); Median (25%-75%)
[2] BMI categories for index cases and household members <18 years of age were defined as BMI z-score (4); 2<, no obesity (underweight, normal weight, overweight); ≥2, obesity. BMI categories for index cases and household members ≥18 years of age were defined as <29.9 kg/m2, no obesity ; ≥30.0 kg/m2, obesity.
[3] Previous infection and vaccination (at least 1 vaccination)

**Table 1b: Baseline characteristics index cases**

| **Characteristic** | **Overall**, N = 145[1] | **Study** | |
|---|---|---|---|
| | | **Sarsliva 1.0**, N = 79[1] | **Sarsliva 2.0**, N = 54[1] |
| **Sex** | | | |
| Male | 44 (33%) | 27 (34%) | 17 (31%) |
| Female | 89 (67%) | 52 (66%) | 37 (69%) |
| **Age, years** | 39 (18-48) | 39 (22-48) | 38 (13-48) |
| **Child/adult** | | | |
| Adult (18 years and older) | 100 (75%) | 63 (80%) | 37 (69%) |
| Child (under 18 years old) | 33 (25%) | 16 (20%) | 17 (31%) |
| **Age group** | | | |
| 0 to 11 years old | 14 (11%) | 1 (1.3%) | 13 (24%) |
| 12 to 17 years old | 19 (14%) | 15 (19%) | 4 (7.4%) |
| 18 to 39 years old | 37 (28%) | 25 (32%) | 12 (22%) |
| 40-49 years old | 38 (29%) | 22 (28%) | 16 (30%) |
| 50 years and older | 25 (19%) | 16 (20%) | 9 (17%) |
| **Disease severity** | | | |
| Asymptomatic | 5 (3.8%) | 5 (6.3%) | 0 (0%) |
| Mild symptoms | 89 (67%) | 52 (66%) | 37 (69%) |
| Moderately severe symptoms | 39 (29%) | 22 (28%) | 17 (31%) |
| **Weight [2]** | | | |
| Obesity | 15 (11%) | 8 (10%) | 7 (13%) |
| No obesity | 130 (90%) | 71 (90%) | 47 (87%) |
| **Any comorbidities** | 50 (38%) | 26 (33%) | 24 (44%) |
| **Atopy** | 41 (31%) | 20 (25%) | 21 (39%) |

| | | | |
|---|---|---|---|
| **Cardiac comorbidities** | 3 (2.3%) | 2 (2.5%) | 1 (1.9%) |
| **Pulmonary comorbidities** | 1 (0.8%) | 1 (1.3%) | 0 (0%) |
| **Immunological comorbidities** | 2 (1.5%) | 1 (1.3%) | 1 (1.9%) |
| **Diabetes Mellitus** | 1 (0.8%) | 0 (0%) | 1 (1.9%) |
| **Rheumatologic comorbidities** | 0 (0%) | 0 (0%) | 0 (0%) |
| **Other comorbidities** | 5 (3.8%) | 3 (3.8%) | 2 (3.7%) |
| **Smoking** | 5 (3.8%) | 3 (3.8%) | 2 (3.7%) |
| **Immunity status** | | | |
| No immunity | 86 (65%) | 79 (100%) | 7 (13%) |
| Previous infection | 5 (3.8%) | 0 (0%) | 5 (9.3%) |
| Vaccination | 38 (29%) | 0 (0%) | 38 (70%) |
| Hybrid immunity [3] | 4 (3.0%) | 0 (0%) | 4 (7.4%) |
| **Antibody titer day 1** | | | |
| **<10** | 3 (6.0%) | - | 3 (6.0%) |
| **10-100** | 0 (0%) | - | 0 (0%) |
| **100-1000** | 5 (10%) | - | 5 (10%) |
| **1000-10.000** | 30 (60%) | - | 30 (60%) |
| **>10.000** | 12 (24%) | - | 12 (24%) |
| Unknown | 83 | 79 | 4 |
| **Antibody titer day 42** | | | |
| **<10** | 5 (4.0%) | 4 (5.4%) | 1 (2.0%) |
| **10-100** | 2 (1.6%) | 2 (2.7%) | 0 (0%) |
| **100-1000** | 45 (36%) | 43 (58%) | 2 (4.0%) |
| **1000-10.000** | 39 (31%) | 25 (34%) | 14 (28%) |
| **>10.000** | 33 (27%) | 0 (0%) | 33 (66%) |
| Unknown | 9 | 5 | 4 |
| **Time from inclusion to last vaccination (days)** | 89 (74-115) | - | 89 (74-115) |
| Unknown | 91 | 79 | 12 |
| **Time from inclusion to last infection (days)** | 118 (96-276) | - | 118 (96-276) |
| Unknown | 124 | 79 | 45 |
| **Time from inclusion to last vaccination or exposure (days)** | 90 (76-116) | - | 90 (76-116) |
| Unknown | 86 | 79 | 7 |
| **Time from symptom onset to GGD-test (days)** | 1 (1-2) | 1 (1-2) | 1 (1-2) |
| **Time GGD-test to saliva day 1 (days)** | 3 (2-4) | 4 (3-4) | 1 (1-2) |
| **Time symptom onset tot inclusion (days)** | 4 (3-5) | 5 (4-5) | 3 (2-4) |

[1] n (%); Median (25%-75%)
[2] BMI categories for index cases and household members <18 years of age were defined as BMI z-score (4); 2<, no obesity (underweight, normal weight, overweight); ≥2, obesity. BMI categories for index cases and household members ≥18 years of age were defined as <29.9 kg/m2, no obesity ; ≥30.0 kg/m2, obesity.
[3] Previous infection and vaccination (at least 1 vaccination)

**Supplementary Table 2. Model setup and outcome.** Table indicates the different types of model, using various input values. We made a division between the cohorts and indicated covariates of each model. The effects of the models are indicated in the last column. A non-trivial effect has a 95%-credibility interval that does not include 0. Significant effects are indicated in bold.

| **Model description** | **cohort** | **Number of input samples** | **Main variable** | **covariates** | **Posterior distribution median (CI 95% lower – upper)** |
|---|---|---|---|---|---|
| **Type 1: shedding decay model** | SL1 | 2131 | Ct value for day since onset | | |
| | SL2 | 1300 | Ct value for day since onset | | |
| **Type 2: ANOVA for minimum Ct value** | SL1 | 213 | Minimum Ct value per participant | Sex | -0.06 (-0.19 – 0.07) |
| | | | | age category (child/adult) | -0.15 (-0.30 – 0.00) |
| | | | | weight class (obesity/ no obesity) | -0.18 (-0.52 – 0.14) |
| | | | | disease severity (category 1 – 4) | **0.14 (0.03 – 0.24)** |
| | | | | transformed SARS-CoV-2 specific antibody titer at day 42 | **0.11 (0.03 – 0.20)** |
| | SL2 | 130 | Minimum Ct value per participant | Sex | -0.04 (-0.21 – 0.13) |
| | | | | age category (child/adult) | **-0.31 (-0.55 – -0.06)** |
| | | | | weight class (obesity/ no obesity) | 0.03 (-0.16 – 0.23) |
| | | | | disease severity (category 1 – 4) | 0.01 (-0.14 – 0.15) |
| | | | | transformed SARS-CoV-2 specific antibody titer at baseline | **-0.13 (-0.26 – 0.00)** |
| | | | | transformed SARS-CoV-2 specific antibody titer at day 42 | **0.18 (0.01 – 0.34)** |
| **Type 3: ANOVA for shedding duration** | SL1 | 210 | Duration of shedding per participant | Sex | **0.01 (0.00 – 0.02)** |
| | | | | age category (child/adult) | -0.00 (-0.01- 0.01) |
| | | | | weight class (obesity/ no obesity) | -0.00 (-0.02 – 0.02) |
| | | | | disease severity (category 1 – 4) | **0.01 (0.00 – 0.02)** |
| | | | | transformed SARS-CoV-2 specific antibody titer at day 42 | **0.01 (0.01 – 0.02)** |
| | SL2 | 130 | Duration of shedding per participant | Sex | -0.00 (-0.01 – 0.00) |
| | | | | age category (child/adult) | **-0.02 (-0.03 - -0.01)** |
| | | | | weight class (obesity/ no obesity) | 0.00 (-0.01 - 0.01) |
| | | | | disease severity (category 1 – 4) | 0.00 (-0.00 – 0.01) |
| | | | | transformed SARS-CoV-2 specific antibody titer at baseline | **-0.01 (-0.02 – -0.00)** |

| | | | | | |
|---|---|---|---|---|---|
| | | | | transformed SARS-CoV-2 specific antibody titer at day 42 | 0.00 (-0.00 – 0.01) |
| *Abbreviations: CI (credibility interval)* | | | | | |
